# Temporal Modeling of Amyloid and Tau Trajectories in Alzheimer’s Disease using PET and Plasma Biomarkers

**DOI:** 10.1101/2025.09.04.25334935

**Authors:** Christopher A. Brown, Katheryn A.Q. Cousins, Magdalena Korecka, Emily McGrew, Alice Chen-Plotkin, John A. Detre, Corey T. McMillan, Edward B. Lee, Sandhitsu R. Das, Dawn Mechanic-Hamilton, Paul A. Yushkevich, Ilya M. Nasrallah, Leslie M. Shaw, the Alzheimer’s Disease Neuroimaging Initiative, David A. Wolk

## Abstract

**Objective:** To compare PET and plasma-based temporal modeling of amyloid and tau biomarkers in Alzheimer’s disease

**Methods:** Longitudinal amyloid PET, ^18^F-flortaucipir tau-PET, and Fujirebio Lumipulse plasma p-tau_217_ from the Alzheimer’s Disease Neuroimaging Initiative (ADNI) and University of Pennsylvania Alzheimer’s Disease Research Center (Penn ADRC) were used to generate biomarker trajectory models using Sampled Iterative Local Approximation (SILA). SILA models using plasma p-tau_217_ were compared to amyloid and tau PET-based models to estimate tau onset age (ETOA) and estimate amyloid onset age (EAOA), and factors influencing ETOA and time from ETOA to dementia were evaluated for PET and plasma-based models.

**Results:** Plasma-based models generated similar results to PET for EAOA and ETOA, with stronger model agreement for ETOA than EAOA. Accuracy of estimated onset age compared to actual onset age was high within modality with slightly greater error when comparing across modalities (i.e. plasma to PET). For both plasma and PET models, earlier ETOA was associated with younger EAOA, female sex, and ≥1 ApoE ε4 allele. Earlier dementia onset after ETOA was associated with later ETOA for both plasma and PET models, while male sex was associated with shorter tau to dementia gap in plasma models.

**Interpretation:** Temporal modeling of plasma biomarkers provides comparable information to PET-based models, particularly for tau onset age. Plasma-based temporal modeling can serve as a widely accessible tool for clinical assessment of biological disease duration that places the patient on the disease timeline, which may allow for improved discussion of prognosis and treatment decisions.

## INTRODUCTION

Alzheimer’s disease (AD) is characterized by the presence of amyloid-β (Aβ) plaques followed by the development of tau neurofibrillary tangles, neurodegeneration, and cognitive decline^1–3^. While the sequence of these events within AD have been well-characterized, there is significant heterogeneity in the specific timing of each of these events^4–6^. Moreover, the presence of co-pathology can alter the timing of neurodegeneration and cognitive impairment relative to amyloid and tau onset^6–9^. Understanding the factors that influence the timing of biomarker changes and alterations to the sequence of these events is particularly important for assessing response to interventions in clinical trials and clinical practice.

Recently developed temporal modeling methods of AD biomarkers allow for estimates of the time of amyloid and tau onset^10^. To date, this literature has focused on amyloid and tau PET to model estimated amyloid and tau onset age (EAOA and ETOA, respectively)^11,12^. After determining these estimated ages, it is possible to anchor longitudinal and cross-sectional analyses to EAOA or ETOA, rather than the arbitrary times at which biomarkers are collected. Using this approach, studies have demonstrated the impact of ApoE status, age, sex, and education on progression to dementia after EAOA, as well as factors influencing ETOA^10–12^. By providing estimated biological disease onset and duration, these methods may be particularly useful in the clinic to improve prognostic predictions, where biomarkers are often not collected until after the onset of symptoms and longitudinal measures are limited by coverage restrictions. While these studies highlight the value of temporal modeling, they have primarily been applied to PET biomarkers, which are less available in clinical settings outside of resource-rich areas.

Over the last several years, significant advances have been made in the accuracy and robustness of plasma biomarkers of AD pathology, especially plasma phosphorylated tau 217 (p-tau_217_) which is strongly associated with both tau and amyloid pathology. This has led to the recent FDA-approval of the first blood-based biomarker for detection of Aβ positivity (Aβ+). As these measures become more ubiquitous in both research and clinical practice, application of temporal modeling approaches to plasma biomarkers could allow for more widespread and direct translation to clinical practice. Here, we used longitudinal data from the Alzheimer’s Disease Neuroimaging Initiative (ADNI) and Penn Alzheimer’s Disease Research Center (ADRC) cohort to generate temporal models of PET and plasma biomarker trajectories using the previously validated sampled-iterative local approximation (SILA) method^10^. We examined the overlap between PET and plasma models, as well as investigated the accuracy of these models in predicting true Aβ and tau positivity in a subset of individuals who converted during longitudinal follow-up. Finally, we examined factors contributing to ETOA and time from ETOA to dementia onset for both PET- and plasma-based models.

## MATERIALS AND METHODS

### Participants

Individuals were selected from ADNI and Penn ADRC based on availability of: 1) Amyloid PET Centiloid (CL) quantification (ADNI only), 2) ^18^F-Flortaucipir tau PET with Tau-MaX calculation^13^, 3) or Fujirebio Lumipulse p-tau_217_. The inclusion and exclusion criteria of these cohorts have been previously published^14,15^, and written informed consent was obtained from all participants under protocols approved by the local Institutional Review Boards (IRBs) in accordance with the Declaration of Helsinki. For both ADNI and Penn ADRC, demographic information and longitudinal clinical diagnoses are collected using procedures described previously^14,15^.

### PET Acquisition and Analysis

Amyloid PET was collected 50-70 minutes or 90-110 minutes after injection using ^18^F-Florbetapir or ^18^F-Florbetaben PET, respectively. For ADNI, images were processed by the ADNI PET core at UC Berkeley to calculate CLs as previously described^16,17^. For Penn ADRC, visual reads of positivity/negativity were performed by an experienced nuclear medicine physician/neuroradiologist (I.M.N). Tau PET was collected 75-105 minutes after injection of ^18^F-Flortaucipir. For ADNI, processed images with uniform 6mm full-width-at-half-maximum resolution were downloaded from the ADNI archive (“Coreg, Av, Std Img and Vox Size, Uniform Resolution”). For Penn ADRC, an in-house pipeline was used to replicate ADNI processing steps as previously described^13^. Pre-processed Tau PET data was then used to calculate global Tau-MaX for all participants as previously described^13^. Briefly, Tau-MaX provides a single global measure of tau burden encompassing both magnitude and extent of tau deposition using a Gaussian mixture-model approach with values ranging between 0-100 with greater values indicating higher tau burden^13^.

### Plasma collection and analysis

Samples were collected as previously described for ADNI and Penn ADRC^18,19^. All samples were analyzed using the Fujirebio Lumipulse G1200 analyzer at either the University of Pennsylvania ADNI core biomarker laboratory (n = 1927 samples) or University of Indiana Biomarker Assay Laboratory (n = 1154 samples) to measure concentrations of p-tau_217_.

### Sampled Iterative Local Approximation (SILA)

We generated separate biomarker trajectories for PET and plasma biomarkers of amyloid and tau using the *silaR* implementation of SILA in R version 4.5.1^10^. For the amyloid PET model, all measurements from participants with at least 2 scans were included and the threshold for positivity was set to CL > 24.4 based on prior post-mortem validation^20^. For the tau PET model, all measurements from Aβ+ participants (CL > 24.4 or positive visual read) with at least 2 Tau-MaX measurements were included, and the threshold for positivity was set to Tau-MaX > 3.31, which represents the 97.5^th^ percentile of cognitively unimpaired (CU) Aβ negative (Aβ−) participants from our prior dataset^13^. For the amyloid and tau plasma model, p-tau_217_ measures were used. For plasma amyloid models, all participants with at least 2 p-tau_217_ measurements available were included, and the Aβ positive threshold of p-tau_217_ > 0.176 based on the maximum Youden index in the combined plasma-PET dataset. For plasma tau models, all Aβ+ participants (p-tau_217_ > 0.176) with at least 2 p-tau_217_ measures were included, and the tau positive threshold of p-tau_217_ > 0.3175 based on maximum Youden index for identifying individuals with Tau-MaX > 3.31 in the combined plasma-PET dataset.

Prior to running SILA models, data were windsorised at CL > 200 = 200, Tau-MaX > 90 = 90, and p-tau_217_ > 2.5 = 2.5, which represented approximate values at which measures were equally likely to go back down rather than continue to rise. SILA models were then run using an integration step-size of 1 year with 100 maximum iterations. We then used these trajectories to estimate amyloid and tau onset age for all participants with at least one measure for a given modality using values from the last available time point with 3 years of extrapolation. This resulted in four estimated onset ages: 1) EAOA_PET_, 2) EAOA_Plasma_, 3) ETOA_PET_, and 4) ETOA_Plasma_.

### Statistical Analysis

R version 4.5.1 was used for all statistical analyses. False discovery rate (FDR) was used for multiple comparison correction with corrected *p* < 0.05 considered significant.

#### Calculating actual onset ages

Actual amyloid onset age (AOA) and tau onset age (TOA) were calculated in a subset of participants who converted from biomarker negative to biomarker positive during longitudinal follow-up. The midpoint between the final biomarker negative timepoint and the first biomarker positive timepoint was considered the actual onset age.

#### Comparing Plasma and PET-based models overlap and accuracy

Correlation analyses evaluated the association between EAOA_PET_-EAOA_Plasma_ and ETOA_PET_-ETOA_Plasma_ across all participants and within Aβ+ individuals. Next, linear regression evaluated EAOA and ETOA predicting of actual AOA and TOA, respectively. Mean absolute error (MAE) was calculated for each regression to assess prediction accuracy. Mean and absolute difference between SILA-estimated onset ages and actual onset ages were also calculated for all measures and comparisons were performed using independent samples t-tests with unequal variance.

#### Examining Factors Influencing ETOA and ETOA-Dementia Gap

We next examined factors influencing ETOA in Aβ+ individuals using Cox-proportional hazards models for plasma and PET models separately. We tested both univariable and multivariable models to evaluate the influence of EAOA, sex, ApoE status, and education on ETOA. ApoE status was divided into two groups based on the presence or absence of ≥ 1 ε4 allele, and education was divided into three groups: high school (HS) with ≤ 12 years, College > 12 but ≤ 16, and Graduate (Grad) with > 16. Finally, we investigated factors influencing time from ETOA to dementia onset in Tau+ individuals. For all individuals who did not have a diagnosis of dementia at their initial visit, time from ETOA was calculated for every visit and time of dementia was considered to be the first time of dementia diagnosis or right censored at time of last follow-up visit. Both univariable and multivariable Cox-proportional hazards models were used for plasma and PET models separately to test the influence of ETOA, sex, ApoE status, and education on ETOA to Dementia time.

## RESULTS

Overall, there were 1733 individuals with CL available, 1087 with Tau-MaX available (ADNI = 875, Penn ADRC = 212), and 1654 individuals with plasma p-tau_217_ (ADNI = 1322, Penn ADRC = 424). There were a total of 1096 individuals with longitudinal CL available with mean 3.05 ± 1.26 amyloid PET scans over 4.77 ± 2.95 years. There were 230 Aβ+ (CL > 24.4) individuals with longitudinal Tau-MaX available with mean 2.85 ± 0.92 tau PET scans over 3.32 ± 1.94 years. For plasma, there were 752 total individuals with longitudinal data available with mean 2.74 ± 0.71 samples over 5.06 ± 2.77 years, while 319 Aβ+ (p-tau_217_ > 0.176) individuals had longitudinal data with mean 2.56 ± 0.72 samples over 4.33 ± 2.69 years. The demographics for the overall and longitudinal (developmental) samples are shown in Table 1.

**Table 1.** Cohort Characteristics.

| SILA Development Dataset |  |  |  |  | Full Dataset |  |  |
| --- | --- | --- | --- | --- | --- | --- | --- |
|  | Amyloid PET<br>n=1096 | Tau PET<br>n=230 | Plasma Amyloid<br>n=752 | Plasma Tau<br>n=319 | Amyloid PET<br>n=1744 | Tau PET<br>n=1086 | Plasma Cohort<br>n=1654 |
| **Age** | 72.5<br>(7.38) | 74.3<br>(7.18) | 72.8<br>(6.93) | 75.3<br>(7.12) | 75.8<br>(8.08) | 74.8<br>(7.79) | 75.4<br>(7.87) |
| **Sex** |  |  |  |  |  |  |  |
| **F** | 533<br>(48.6%) | 120<br>(52.2%) | 366<br>(48.7%) | 145<br>(45.5%) | 851<br>(49.1%) | 599<br>(55.2%) | 857<br>(51.8%) |
| **M** | 563<br>(51.4%) | 110<br>(47.8%) | 386<br>(51.3%) | 174<br>(54.5%) | 883<br>(50.9%) | 487<br>(44.8%) | 797<br>(48.2%) |
| **Education (Yrs)** | 16.4<br>(2.57) | 16.2<br>(2.38) | 16.3<br>(2.62) | 16.1<br>(2.7) | 16.3<br>(2.57) | 16.4<br>(2.45) | 16.3<br>(2.55) |
| **Amyloid** |  |  |  |  |  |  |  |
| **Positive** | 545<br>(49.7%) | 230<br>(100%) | 287<br>(38.2%) | 319<br>(100%) | 881<br>(49.2%) | 438<br>(42.2%) | 807<br>(48.8%) |
| **Negative** | 551<br>(50.3%) | 0 | 465<br>(61.8%) | 0 | 853<br>(50.8%) | 599<br>(57.8%) | 847<br>(51.2%) |
| **ApoE ε4** |  |  |  |  |  |  |  |
| **Absent** | 614<br>(58.9%) | 80<br>(38.8%) | 362<br>(62.0%) | 127<br>(46.5%) | 892<br>(57%) | 437<br>(60.7%) | 626<br>(57.3%) |
| **Present** | 429<br>(41.1%) | 126<br>(61.2%) | 222<br>(38.0%) | 146<br>(53.5%) | 672<br>(43%) | 283<br>(39.3%) | 468<br>(42.7%) |
| **Diagnosis** |  |  |  |  |  |  |  |
| **CU** | 508<br>(48.3%) | 111<br>(48.7%) | 395<br>(56.0%) | 112<br>(37.9%) | 525<br>(35.7%) | 527<br>(54.9%) | 742<br>(50.1%) |
| **MCI** | 471<br>(44.8%) | 75<br>(32.9%) | 259<br>(36.7%) | 135<br>(45.8%) | 592<br>(40.3%) | 287<br>(29.9%) | 454<br>(30.6%) |
| **Dementia** | 72<br>(6.9%) | 42<br>(18.4%) | 51<br>(7.2%) | 48<br>(16.3%) | 352<br>(24%) | 145<br>(15.1%) | 286<br>(19.3%) |

Mean (SD) for continuous measures and count (%) for categorical variables. Age is baseline age for SILA development dataset and age at last time point (used to estimate onset) in full dataset. F: Female, M: Male, CU: Cognitively Unimpaired, MCI: Mild Cognitive Impairment

### SILA Models of Amyloid Trajectory

The amyloid PET SILA trajectory and ΔCL by CL curves are shown in Figure 1A. Average EAOA_PET_ was 76.6 ± 15.6 years for all individuals and 66.8 ± 9.48 years for Aβ+ individuals. The plasma amyloid SILA trajectory and Δp-tau_217_ by p-tau_217_ curves are shown in Figure 1B. Average EAOA_Plasma_ was 74.7 ± 9.76 years for all individuals and 69.92 ± 9.56 years for Aβ+ individuals. In the 1138 individuals with both CL and p-tau_217_ available, there was a strong correlation between EAOA_PET_ and EAOA_Plasma_ (*r* = 0.72, *df* = 1136, *p* < 0.001), while there was a stronger correlation in the 545 Aβ+ individuals with both CL and p-tau_217_ available (*r* = 0.82, *df* = 543, *p* < 0.001) (Figure 1C). There were 103 individuals in the Amyloid PET dataset, 110 individuals in the plasma dataset, and 94 in the combined dataset that converted from Aβ− to Aβ+ during longitudinal follow-up. The MAE was 1.54 for EAOA_PET_-AOA_PET_, 2.04 for EAOA_Plasma_-AOA_Plasma_, and 3.49 for cross-modal EAOA_Plasma_-AOA_PET_ (Figure 2A). The mean difference between EAOA-AOA did not differ for PET and plasma models (Table 2), but the absolute mean difference was slightly larger for EAOA-AOA for plasma compared to PET (*t* = −2.02, *df* = 182.2, *p* = 0.045).

**Figure 1.**
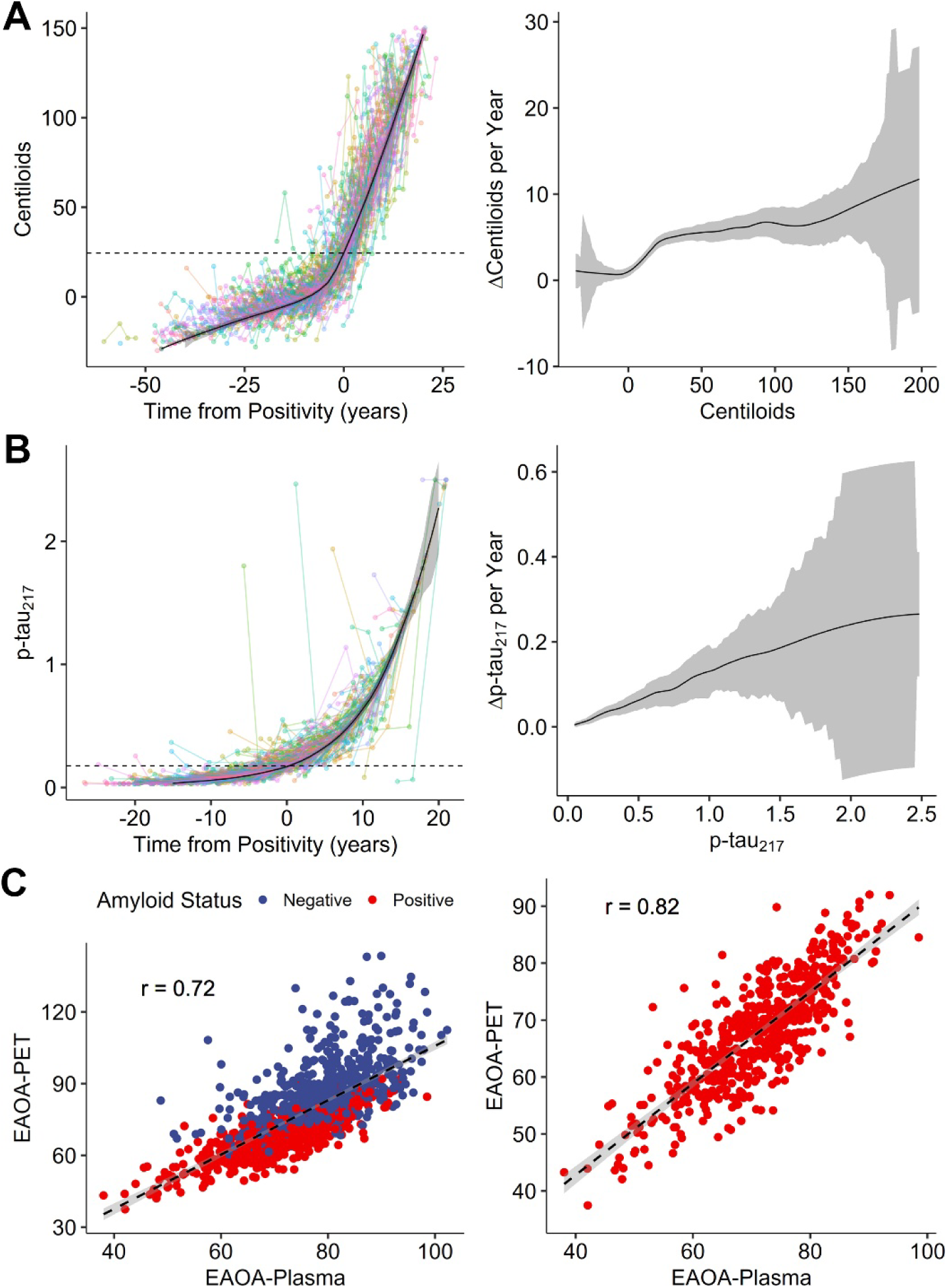
Temporal modeling of amyloid trajectories using PET and plasma. **A-B:** Longitudinal trajectories (left panels) for PET (A) and plasma (B) amyloid with each point representing a single timepoint and lines connecting points from the same participant. Dashed horizontal lines indicate amyloid positivity threshold. SILA trajectory curves are shown with shading representing the error. On the right, estimated rate of change for a given CL (A) or p-tau_217_ (B) value is shown with error shown by the shaded ribbon. **C:** Association between EAOA_PET_ and EAOA_Plasma_ in all individuals (left) and within Aβ+ individuals (right). The dashed line represents the linear best-fit with 95% confidence of fit shown by the shaded ribbon.

**Figure 2.**
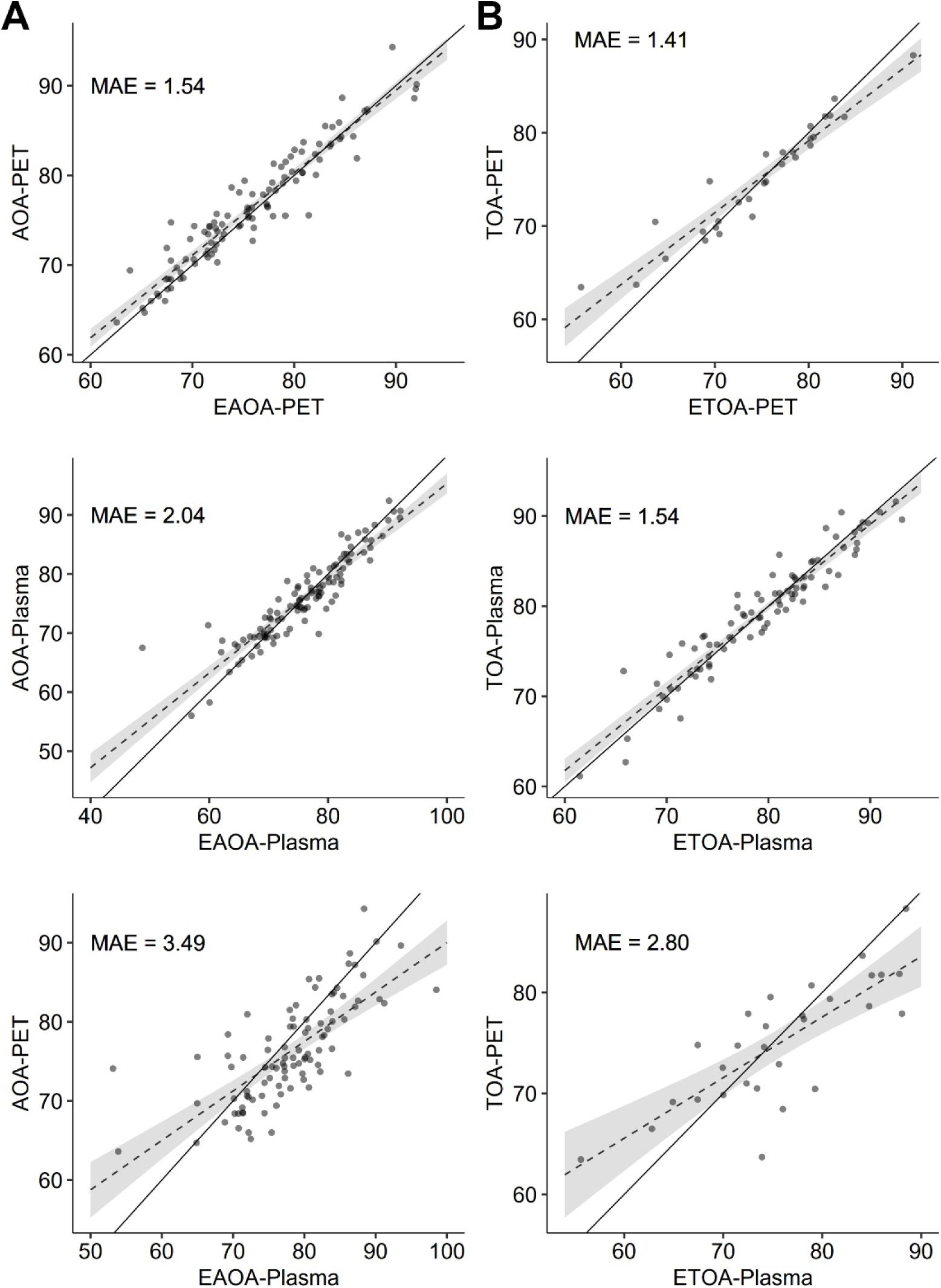
Comparison of estimated and actual onset age. **A:** The association between Estimated Amyloid Onset Age (EAOA) and actual Amyloid Onset Age (AOA) for PET (top), plasma (middle) and EAOA_Plasma_ for AOA_PET_ (bottom) in the subset of individuals who converted from negative to positive during follow-up. **B:** The association between Estimated Tau Onset Age (ETOA) and actual Tau Onset Age (TOA) for PET (top), plasma (middle) and ETOA_Plasma_ for TOA_PET_ (bottom) in the subset of individuals who converted from negative to positive during follow-up. **A-B:** The dashed line represents the linear best-fit with the ribbon showing the 95% confidence of fit, while the solid line is the identity line shown for reference (slope=1). MAE: Mean absolute error

**Table 2.** SILA Models Prediction Error for Actual Onset Age.

|  | EAOA<sub>PET</sub> | EAOA<sub>Plasma</sub> | ETOA<sub>PET</sub> | ETOA<sub>Plasma</sub> | Plasma-PET Comparison |  |
| --- | --- | --- | --- | --- | --- | --- |
|  |  |  |  |  | Amyloid | Tau |
| **MAE** | 1.54 | 2.04 | 1.41 | 1.54 | -- | -- |
| **Cross-Modal MAE** | -- | 3.49 | -- | 2.80 | -- | -- |
| **Mean Difference** | 0.63 | 0.10 | 0.36 | 0.05 | $t(187) = 1.45$<br>$p = 0.15$ | $t(41.8) = 0.61$<br>$p = 0.54$ |
| **Mean Absolute Difference** | 1.56 | 2.11 | 1.62 | 1.54 | $t(182) = 2.02$<br>$p = 0.045$ | $t(38.0) = 0.21$<br>$p = 0.84$ |

T-tests comparing error of PET and plasma models for amyloid and tau; MAE: Mean absolute error

### SILA Models of Tau Trajectory

The tau PET SILA trajectory and ΔTau-MaX by Tau-MaX curves are shown in Figure 3A. Average ETOA_PET_ was 76.8 ± 8.59 years for all individuals, 76.1 ± 9.97 years for Aβ+ individuals, and 71.4 ± 9.53 years for Tau+ (Tau-MaX > 3.31) individuals. The plasma tau SILA trajectory and Δp-tau_217_ by p-tau_217_ curves are shown in Figure 3B. Average ETOA_Plasma_ was 77.7 ± 8.87 years across all individuals, 74.8 ± 9.41 for Aβ+ individuals, and 72.2 for Tau+ (p-tau_217_ > 0.3175) individuals. In the 1012 individuals with both Tau-MaX and p-tau_217_ available, there was a strong correlation between ETOA_PET_ and ETOA_Plasma_ (*r* = 0.89, *df* = 1010, *p* < 0.001), while the correlation slightly increased within 378 Aβ+ individuals (*r* = 0.92, *df* = 376, *p* < 0.001) (Figure 3C). There were 29 individuals in the Tau PET dataset, 84 individuals in the plasma dataset, and 29 in the combined dataset that converted from Tau− to Tau+ during longitudinal follow-up. Within this group, the MAE was 1.41 for ETOA_PET_-TOA_PET_, 1.54 for ETOA_Plasma_-TOA_Plasma_, and 2.80 for the multi-modality ETOA_Plasma_-TOA_PET_ (Figure 2B). Neither the mean or absolute difference between ETOA-TOA differed between PET and plasma models (Table 2).

**Figure 3.**
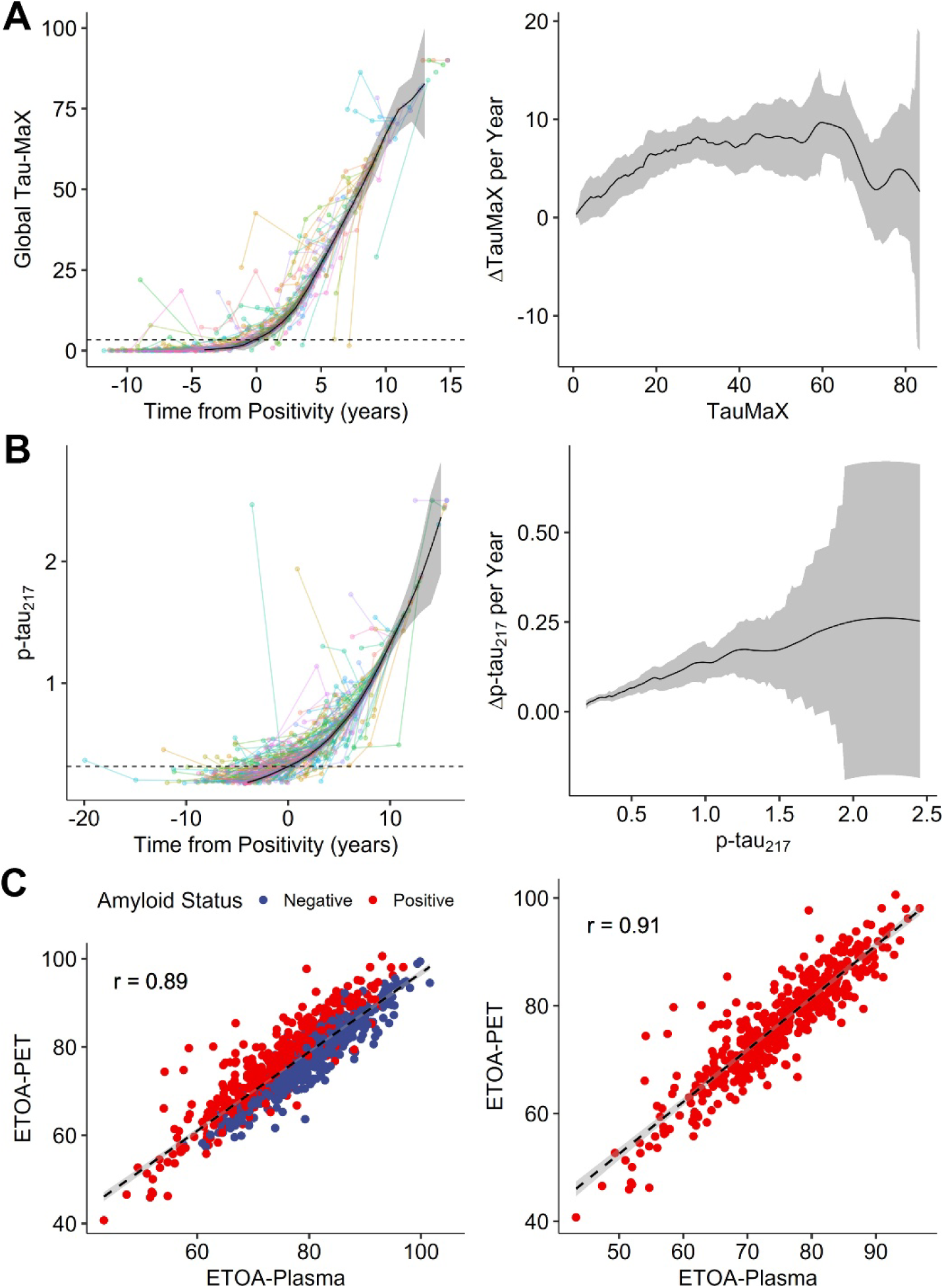
Temporal modeling of tau trajectories using PET and plasma. **A-B:** Longitudinal trajectory (left) for PET (A) and plasma (B) amyloid with each point representing a single timepoint and lines connecting points from the same participant. SILA trajectory curves are shown with shading representing the error. On the right, estimated rate of change for a given Tau-MaX (A) or p-tau_217_ (B) value is shown with error shown by the shaded ribbon. **C:** Association between ETOA_PET_ and ETOA_Plasma_ in all individuals (left) and within Aβ+ individuals (right). The dashed line represents the linear best-fit with 95% confidence of fit shown by the shaded ribbon.

### Factors Influencing ETOA

We first examined univariable models to assess factors influencing ETOA_PET_ and ETOA_Plasma_ separately in Aβ+ individuals. For both the PET and plasma models, younger ETOA was associated with younger EAOA, female sex, and presence of at least one ApoE ε4 allele (Figure 4A). We then tested multivariable models with all measures included simultaneously and found that all results from univariable models remained significant in the multivariable model, such that earlier EAOA, female sex, and presence of at least one ApoE ε4 allele were all associated with earlier ETOA for both PET and plasma derived measures (Figure 4B).

**Figure 4.**
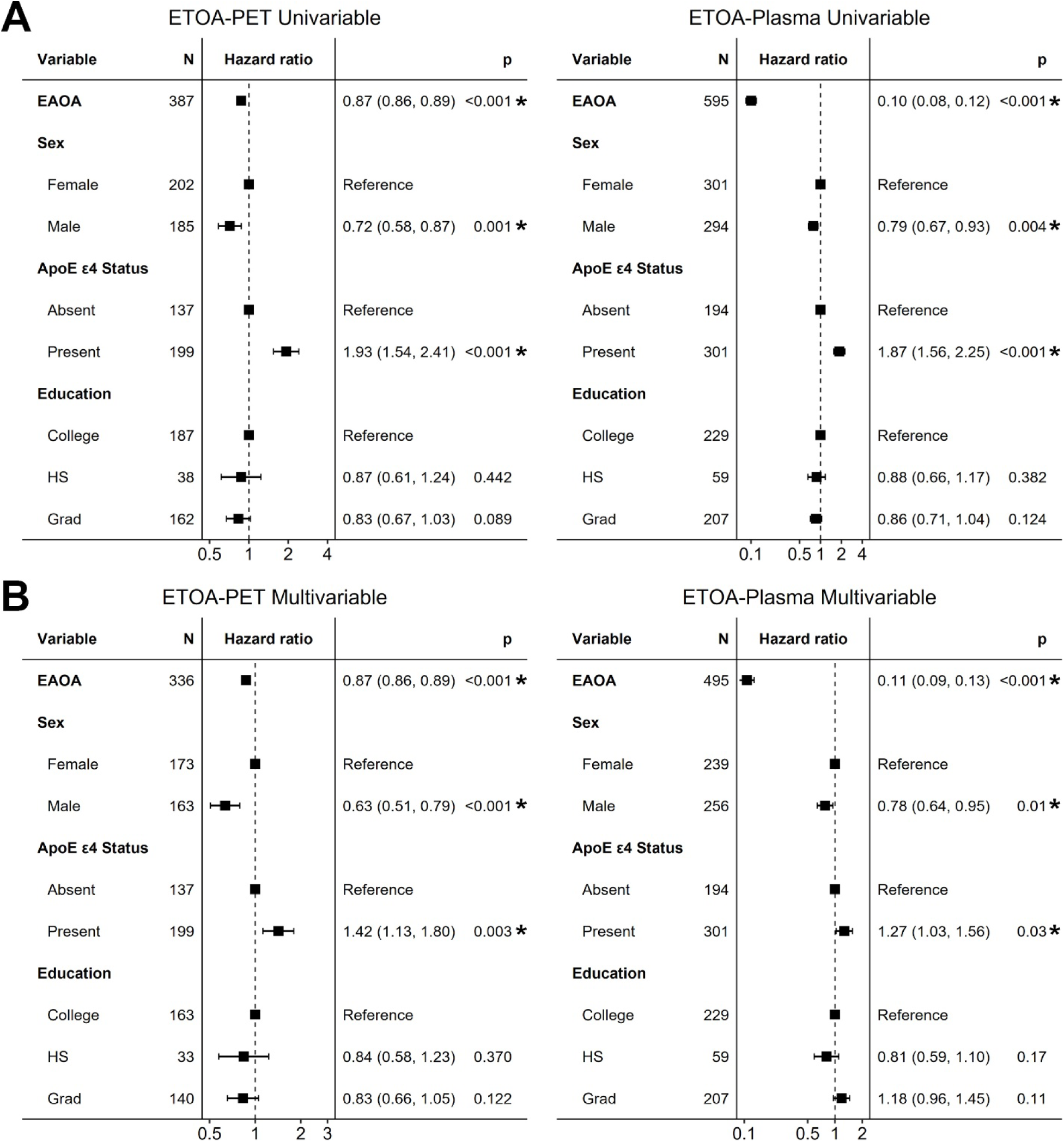
Factors influencing Estimated Tau Onset Age. Univariable (A) and multivariable (B) cox-proportional hazards models examining the effects of various factors on Estimated Tau Onset Age (ETOA) from PET (left) and plasma (right) models. Estimated Amyloid Onset Age (EAOA) is from within-modality. Hazard ratios from Cox-proportional hazards are shown with 95% confidence intervals and uncorrected p-values. \**p_FDR_*< 0.05.

### Factors Influencing ETOA to Dementia Gap

Next, we evaluated the factors influencing time from ETOA-to-dementia onset in Tau+ individuals who did not have a diagnosis of dementia at their baseline visit. In the Tau-PET cohort 124/245 Tau+ individuals transitioned to dementia during follow-up, while in the p-tau_217_ cohort 306/541 Tau+ individuals transitioned. Univariable models found that younger ETOA was associated with longer ETOA-to-dementia gap in both the PET and Plasma models, while female sex was also associated with longer ETOA to dementia gap in the plasma model (Figure 5A). For the PET model, similar results were seen in the multivariable model, with only earlier ETOA_PET_ associated with longer ETOA to dementia gap (Figure 5B). In contrast, for the plasma model both earlier ETOA_Plasma_ and female sex were associated with longer dementia-free survival after ETOA, while there was a trend towards shorter ETOA to dementia gap in individuals with at least one ApoE ε4 allele and longer ETOA to dementia gap in individuals with graduate education (Figure 5B).

**Figure 5.**
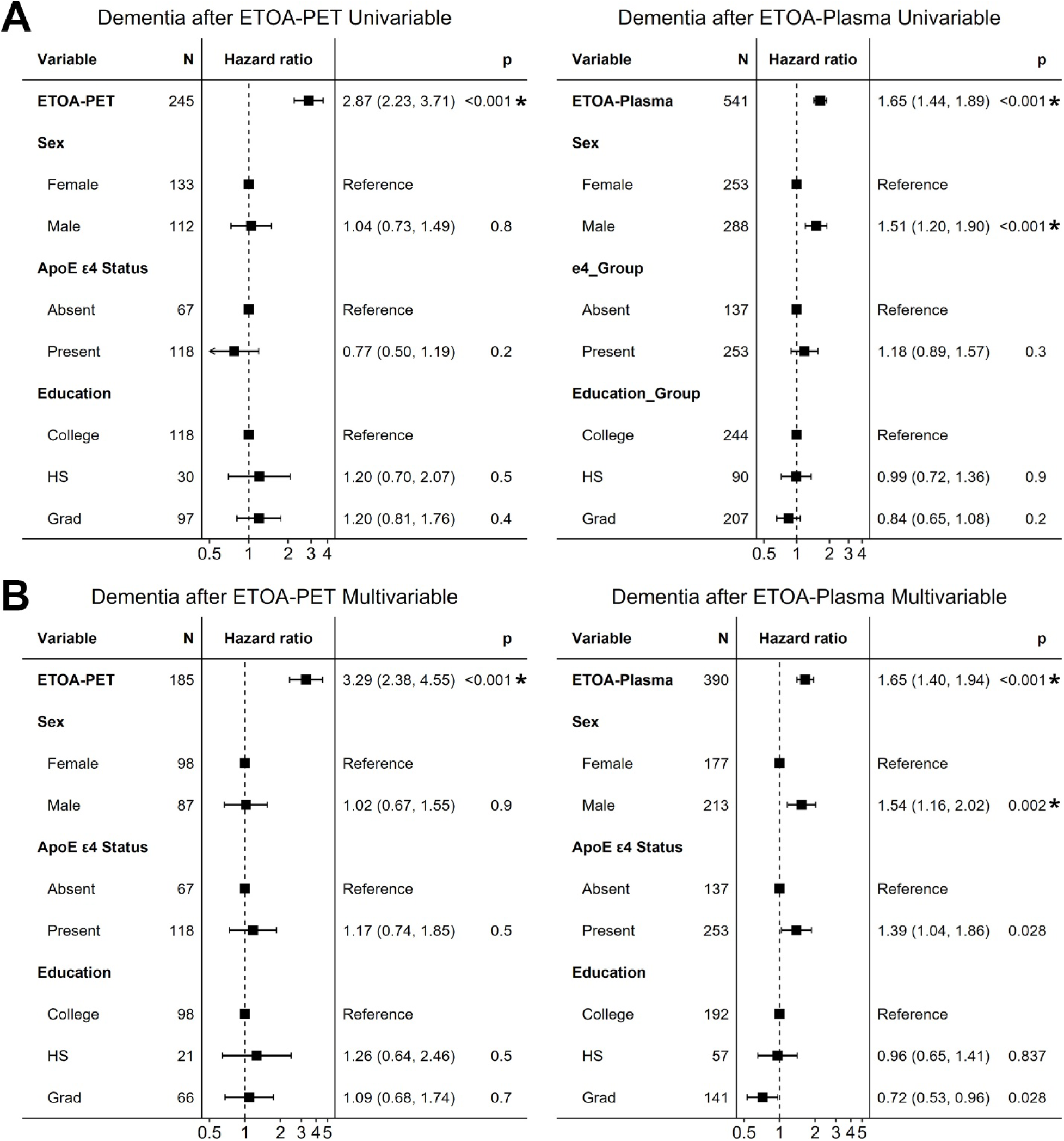
Factors influencing Time from Estimated Tau Onset Age to Dementia. Univariable (A) and multivariable (B) cox-proportional hazards models examining the effects of various factors on time from Estimated Tau Onset Age (ETOA) from PET (left) and plasma (right) to dementia diagnosis. To allow for comparability to other factors, ETOA is scaled to represent the risk of every 10 year change. Hazard ratios from Cox-proportional hazards are shown with 95% confidence intervals and uncorrected p-values. \**p_FDR_* < 0.05.

## DISCUSSION

Our study extends prior temporal modeling techniques for both amyloid and tau trajectories to plasma-based models and demonstrates comparability between these two modalities. There was high overlap for both EAOA and ETOA between PET and plasma models, although there was slightly higher agreement in tau models and no difference in error for predicting TOA between modalities. Further, we found largely overlapping factors that influenced ETOA and time from ETOA to dementia between PET- and plasma-based models. Overall, our findings demonstrate that plasma-based temporal modeling of amyloid and tau trajectories place individuals on the biological timeline of AD with minimal loss compared to PET-based estimates and, thus, provide a highly accessible alternative approach.

Our amyloid PET model is similar to recently developed SILA models using CL, although we opted for a higher CL cutoff of 24.4 based on prior post-mortem work and real-world data that suggest that these higher CL cutoffs are closely aligned with the presence of AD pathology at autopsy and PET visual reads^20,21^. As a result, the mean EAOA_PET_ in this study is very similar to that previously identified using the largely overlapping ADNI PET data but with a CL > 19 considered abnormal^11^. However, we extend the SILA approach beyond amyloid PET to a global measure of tau burden using Tau-MaX, as well as to a plasma biomarker of amyloid and tau, p-tau_217_. A recent study applied temporal modeling to Tau-PET but used temporal meta-ROI standardized uptake value ratio (SUVR), which is less sensitive to non-canonical patterns of tau accumulation and appears to be more susceptible to ceiling effects, particularly with ^18^F-Flortaucipir^11,13^. These findings further highlight the flexibility of temporal modeling to place individuals on the AD pathological timeline using different biomarkers and modalities.

Importantly, there was high overlap between EAOA and ETOA estimated by PET and plasma biomarkers, particularly within Aβ+ individuals. Further, this association was higher for ETOA than EAOA, consistent with a stronger association between plasma p-tau_217_ and Tau-PET compared to amyloid PET, despite p-tau_217_ frequently being thought of as a marker specifically of amyloid positivity^13,18,22^. When examining average onset age across all measures, we found that biomarkers became abnormal in the following order: 1) Amyloid PET, 2) p-tau_217_ Aβ cutoff, 3) p-tau_217_ tau cutoff, 4) Tau PET, which is consistent with the current understanding of the AD pathological cascade with accumulation of amyloid plaques leading to abnormal phosphorylation of soluble tau species and then formation of insoluble tau neurofibrillary tangles that can be measured with Tau PET^3^. The degree to which each marker represents slightly different events in the neuropathological cascade may also be the primary reason for considerably greater error when attempting to predict onset across modalities despite high correlation.

In addition to high overlap between PET and plasma EAOA and ETOA, we also found that using PET or plasma-based estimates resulted in similar factors influencing ETOA. Specifically, we found that earlier EAOA, female sex, and presence of at least one ApoE ε4 allele were associated with earlier ETOA in both univariable and multivariable models. These findings are consistent with studies showing that early-onset AD is associated with earlier (and greater) tau accumulation compared to late-onset AD^23^, women have higher tau loads for a given level of amyloid compared to men^23,24^, and ApoE plays an important role in tau accumulation that is independent of amyloid ^25,26^. Further, these findings are similar to a prior study using SILA to investigate factors influencing ETOA^11^.

In contrast to factors influencing ETOA, multivariable models investigating the factors associated with dementia-free survival following ETOA showed larger differences between PET and plasma models. While PET models showed that only earlier ETOA was associated with longer dementia free survival after ETOA, plasma models found that both ETOA and sex had significant effects, while education and ApoE status had marginal effects. One previous study has found similar associations between earlier ETOA and dementia-free survival after ETOA using PET^11^. However, our plasma findings are consistent with prior work showing that individuals with early-onset AD tend to have onset of symptoms at much later biological disease stage compared to late-onset cases^23,27^, women have later biological disease stage at symptom onset compared to men^24,28,29^, and those with at least one ApoE ε4 allele have faster cognitive decline than non-carriers^28,30,31^. Similarly, a large body of research in cognitive reserve has demonstrated that factors such as education are associated with higher resilience to tau pathology with later onset of cognitive symptoms^32^. Of note, these findings in the plasma model are likely related to stronger power, as all effects except for ETOA disappear when restricted to an overlap cohort with both tau PET and plasma.

This study has several limitations. First, due to lack of alternative plasma biomarkers, we used the same measure to estimate EAOA and ETOA in blood, thus creating a stronger dependence of ETOA on EAOA. We did alter the samples used to generate these trajectories, but there is still significant overlap between these models. From a biological perspective, p-tau_217_ clearly captures aspects of both amyloid and tau pathology, making it sensitive but less specific for either of these pathologies individually. Development of more specific biomarkers of tau, such as microtubule-binding region containing tau residue 243 (MBTR-tau243), may help in isolating tau-specific and amyloid-specific aspects of p-tau_217_ to further refine these models^33^. Second, our assessment of factors influencing ETOA and time from ETOA to dementia was relatively limited to measures widely available within these datasets. Future work will be necessary to more fully evaluate the influence of various factors, particularly those associated with genetics and social determinants of health, as well as further examination of factors that may contribute to individual error in EAOA and ETOA prediction.

A major strength of this study is the application of temporal modeling to two new domains: 1) global tau burden measured by Tau-PET using Tau-MaX and 2) plasma p-tau_217_. These models provide valuable tools for anchoring the disease course to biological milestones without needing to directly observe these transitions during longitudinal follow-up and move beyond simple dichotomous designations of positive and negative. Further, the estimation of an age of onset rather than raw biomarker measurement values (e.g. SUVR) provides a more conceptually tractable tool for understanding biological stage. This may be of particular value when assessing individuals clinically, who often do not present until after the onset of cognitive symptoms. In this setting, obtaining p-tau_217_ could allow for placing an individual on the amyloid and tau timeline, and, after combining this with their demographics and ApoE status, provide prognostic guidance for likely time to dementia onset. This may be of even greater importance in the era of disease-modifying therapies, where providing a general sense of an individual’s time from ETOA and onset of dementia could be valuable when making treatment decisions. Furthermore, large discrepancies between clinical severity and what would be expected at a given biological disease duration may suggest presence of co-pathology or resilience^8,9,34^, which again may impact treatment decisions.

In conclusion, we generated temporal models of amyloid and tau trajectories for both PET and plasma markers of pathology. These models show high overlap and comparable error, particularly for predicting tau onset. Furthermore, this approach helps to further elucidate the important role of age of amyloid onset, sex, and ApoE status in disease trajectory. Together, these findings highlight the potential value of temporal modeling approaches and their direct ability for translation to clinic via widely collected plasma biomarkers of AD pathology.

## ACKNOWLEDGEMENTS

This work was supported by grants from the National Institutes of Health (P30-AG072979, RF1-AG069474, R01-AG056014, R01-AG055005, R01-AG072796, R25-NS065745, P01-AG084497), Pennsylvania Department of Health (2019NF4100087335), and Alzheimer’s Association and Fred and Barbara Erb Family Foundation (AACSF-23-1152241). Data collection and sharing for the Alzheimer’s Disease Neuroimaging Initiative (ADNI) is funded by the National Institute on Aging (National Institutes of Health Grant U19 AG024904). The grantee organization is the Northern California Institute for Research and Education. In the past, ADNI has also received funding from the National Institute of Biomedical Imaging and Bioengineering, the Canadian Institutes of Health Research, and private sector contributions through the Foundation for the National Institutes of Health (FNIH) including generous contributions from the following: AbbVie; Alzheimer’s Association; Alzheimer’s Drug Discovery Foundation; Araclon Biotech; BioClinica, Inc.; Biogen; Bristol-Myers Squibb Company; CereSpir, Inc.; Cogstate; Eisai Inc.; Elan Pharmaceuticals, Inc.; Eli Lilly and Company; EuroImmun; F. Hoffmann-La Roche Ltd and its affiliated company Genentech, Inc.; Fujirebio; GE Healthcare; IXICO Ltd.; Janssen Alzheimer Immunotherapy Research & Development, LLC; Johnson & Johnson Pharmaceutical Research &Development LLC; Lumosity; Lundbeck; Merck & Co., Inc.; Meso Scale Diagnostics, LLC; NeuroRx Research; Neurotrack Technologies; Novartis Pharmaceuticals Corporation; Pfizer Inc.; Piramal Imaging; Servier; Takeda Pharmaceutical Company; and Transition Therapeutics.

## AUTHOR CONTRIBUTIONS

Conception and design of the study: C.A.B., K.A.Q.C., A.C.P., J.A.D., C.T.M., E.B.L., D.M.H., P.A.Y., I.M.N., L.M.S., D.A.W.

Acquisition and analysis of data: C.A.B., K.A.Q.C., M.K., E.M., S.R.D., D.M.H., I.M.N., L.M.S. Drafting a significant portion of the manuscript or figures: C.A.B.

## DATA AVAILABILITY

All requests for raw and analyzed data from the Penn ADRC cohort will be reviewed by the Penn Neurodegenerative Data Sharing Committee (PNDSC) and shared for appropriate uses through a data sharing agreement (https://www.pennbindlab.com/data-sharing). Anonymized data from Penn ADRC will be shared upon request to the corresponding author by a qualified academic investigator for the purpose of replicating procedures and results in this article. Data are not publicly available due to privacy protections outlined in the participant informed consent. Documents related to study protocols, informed consent and other documentation can similarly be made available upon request. All ADNI data are shared without embargo through the LONI Image and Data Archive (https://ida.loni.usc.edu/), a secure research data repository. Interested scientists may obtain access to ADNI imaging, clinical, genomic, and biomarker data for the purposes of scientific investigation, teaching, or planning clinical research studies. Access is contingent on adherence to the ADNI Data Use Agreement and the publications’ policies (https://adni.loni.usc.edu/data-samples/access-data/).

